# Antigen production and development of an indirect ELISA based on the nucleocapsid protein to detect human SARS-CoV-2 seroconversion

**DOI:** 10.1101/2021.02.19.21252100

**Authors:** Marcelo S. Conzentino, Karl Forchhammer, Emanuel M Souza, Fábio O. Pedrosa, Meri B. Nogueira, Sônia M. Raboni, Fabiane G. M. Rego, Dalila L. Zanette, Mateus N. Aoki, Jeanine M. Nardin, Bruna Fornazari, Hugo M. P. Morales, Paola A. F. Celedon, Carla V. P. Lima, Sibelle B. Mattar, Vanessa H. Lin, Luis G. Morello, Fabricio K. Marchini, Lucas Bochnia Bueno, Rodrigo A. Reis, Luciano F. Huergo

## Abstract

Serological assays are important tools to identify previous exposure to SARS-CoV-2, helping to track COVID-19 cases and determine the level of humoral response to SARS-CoV-2 infections and/or immunization to future vaccines. Here the SARS-CoV-2 nucleocapsid protein was expressed in *Escherichia coli* and purified to homogeneity and high yield using a single chromatography step. The purified SARS-CoV-2 nucleocapsid protein was used to develop an indirect enzyme-linked immunosorbent assay for the identification of human SARS-CoV-2 seroconverts. The assay sensitivity and specificity were determined analyzing sera from 140 PCR-confirmed COVID-19 cases and 210 pre-pandemic controls. The assay operated with 90% sensitivity and 98% specificity; identical accuracies were obtained in head-to-head comparison with a commercial ELISA kit. Antigen coated plates were stable for up to 3 months at 4°C). The ELISA method described is ready to mass production and will be an additional toll to track COVID-19 cases.

## Introduction

In December 2019, health authorities in Wuhan, China, reported cases of patients with pneumonia of unknown epidemiological causes, linked to a seafood/animal market. The pathogen in these cases has been identified, through viral isolation, electron microscopy and RNA sequencing, as a new beta-coronavirus called SARS-CoV-2 (Zhu *et al*., 2020). Shortly after its emergence in China, SARS-CoV-2 spread rapidly across the world. According to the John Hopkins University website (https://coronavirus.jhu.edu/map.html), on 01/02/2021, more than 100 million cases of COVID-19 have been registered worldwide and the disease was responsible for 2,236,000 deaths.

Serological assays are important for epidemiological surveillance studies to monitor COVID-19 cases (Petherick, 2020). The structural Nucleocapsid (N) protein is known to be a major antigen of coronavirus producing a strong humoral response in humans (Cheng *et al*., 2007). Furthermore, the SARS-CoV-2 Nucleocapsid protein can be used as antigen to detect COVID-19 cases without significant cross-reactivity with common cold coronavirus (den Hartog *et al*., 2020). Here we describe the develop an indirect enzyme-linked immunosorbent assay (ELISA) which allowed accurate detection human anti-IgG SARS-CoV-2 seroconversion.

## Results and Discussion

A codon optimized synthetic gene was obtained and cloned into the pET28a vector to allow the expression of an N-terminal 6xHis tagged SARS-CoV-2 N protein in *Escherichia coli* BL21 (λDE3) (Huergo *et al*., 2020; F. Huergo *et al*., 2021). The viral protein was highly expressed and remained partially soluble in *E. coli* cell extracts (Fig. 1A). The protein was purified using a 5 ml His-trap affinity column (Cytiva) followed by elution with an imidazole gradient. The His-tagged SARS-CoV-2 N protein eluted at 300 mM of imidazole to apparent homogeneity on Coomassie stained SDS-PAGE (Fig. 1A). Typical protein yield was ∼8 mg per 100 ml of culture, the antigen was stored for up to 4 months at -20°C.

**Fig 1.**
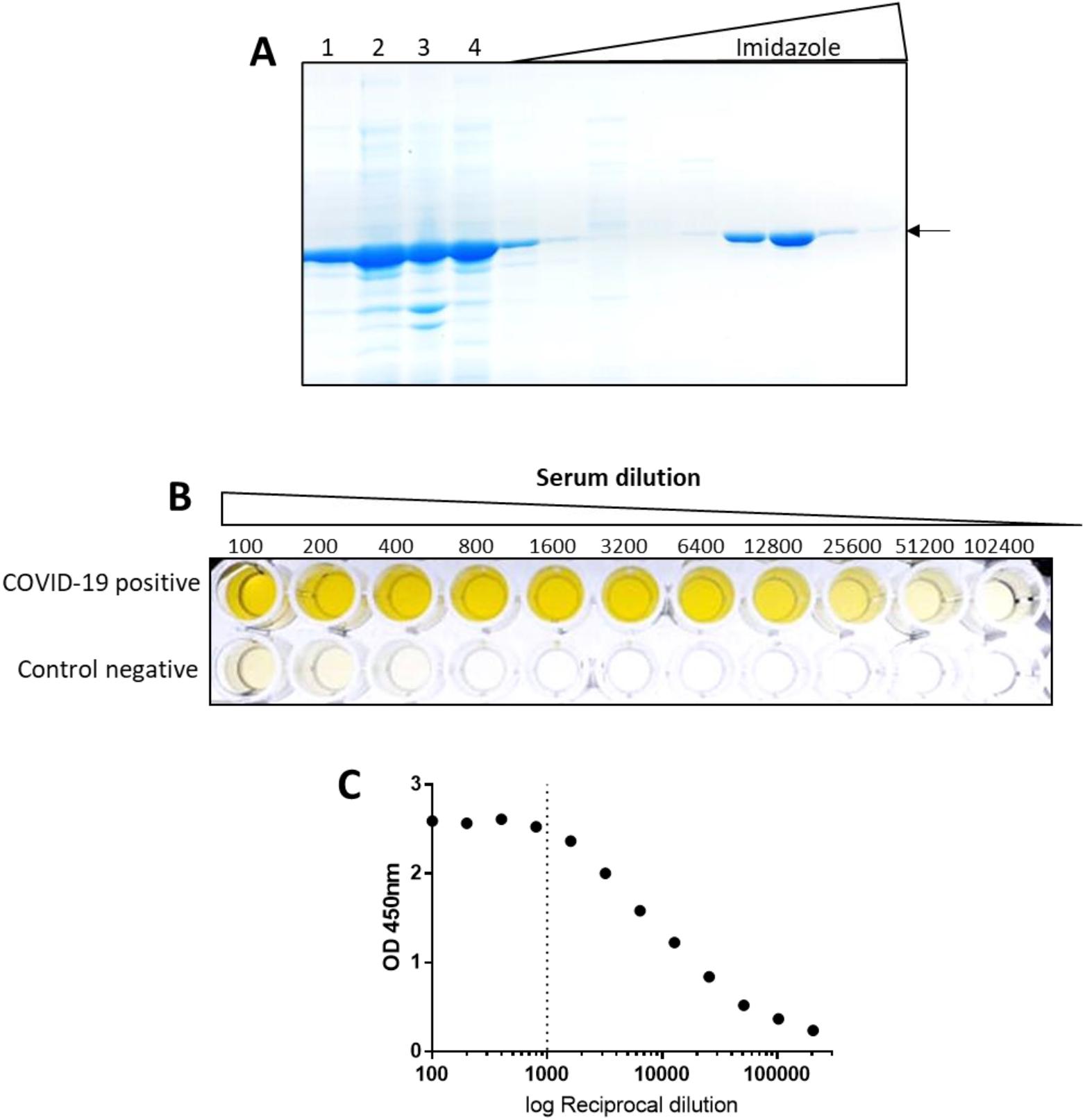
Purification of the SARS-CoV-2 Nucleocapsid protein and serum dilution analysis on ELISA. A) SDS-PAGE analysis of purified His-tagged SARS-CoV-2 N protein. Lane 1, whole cell extract; Lane 2, soluble fraction; Lane 3, insoluble fraction; Lane 4, column flow through; Other lanes, imidazole gradient, the N protein is indicated by an arow and eluted at 300 mM imidazole. B) The serum from a COVID-19 positive case and from a negative control were serial diluted and analyzed using indirect ELISA, the data were recorded photographically. C) The data derived from the COVID-19 positive case was expressed as OD values vs reciprocal dilution; the dashed line indicates the dilution used during this study.

The purified N protein was diluted in buffer and used to coat 96 well polystyrene plates overnight at 4°C. Plates were blocked with skimmed milk and serum from a COVID-19 positive case and a negative control serum were serial diluted and placed into the wells of pre-coated plates. The presence of IgG reacting with the SARS-CoV-2 N protein was revealed by incubating with secondary anti-human IgG -HPR followed by chromogenic substrate TMB. The COVID-19 positive serum showed strong reaction with the N protein even at high dilutions (Fig. 1B). On the other hand, the negative control serum showed only minor background cross reaction in low dilutions (Fig. 2A). The OD450nm response vs serum dilution of the positive serum was linear between the 1,600 to 25,600 dilutions with R^2^ = 0.995 (Fig. 1C). This positive serum was used as a positive control throughout this study, the inter assay CV% was 7.2% (same sample run in 12 different plates in duplicate). The mean intra assay CV% was 7.6% (mean CV% was obtained from duplicate 96 samples x 12 plates).

**Fig 2.**
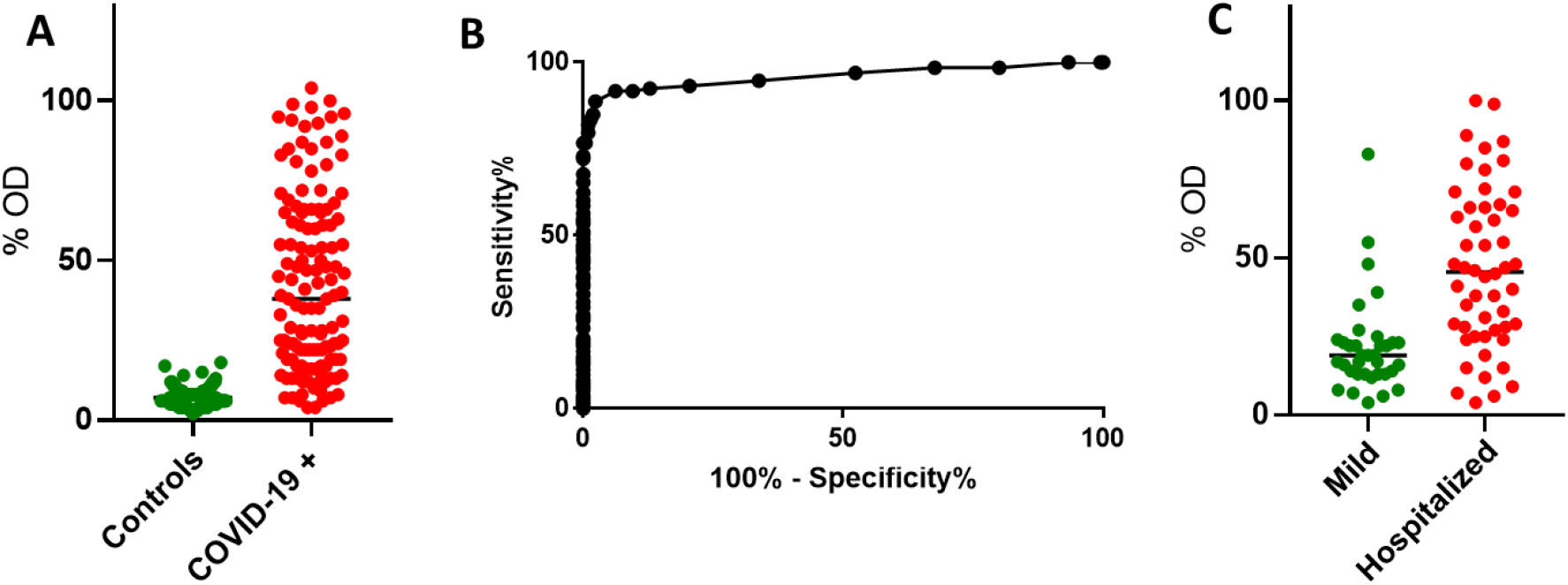
IgG seroconversion response to SARS-CoV-2 infection. A) The OD was expressed as % of reference serum for negative control samples (n=210) and PCR-confirmed COVID-19 cases (n=140). B) Receiver Operating Characteristics analysis (ROC) of the data described in A. C) Comparison between the IgG signal reacting with the SARS-CoV-2 Nucleocapsid protein in Mild and Hospitalized PCR-confirmed COVI-19 cases. Groups were compared using the T test.

## Material and Methods

### Expression and purification of recombinant SARS-CoV-2 Nucleocapsid protein (N-protein)

A codon optimized synthetic gene expressing the full-length SARS-CoV-2 N-protein (Uniprot QHD43423.2) was obtained and cloned into pET28a by General Biosystems (F. Huergo *et al*., 2021). This plasmid was named pLHSarsCoV2-N and transformed into *E. coli* BL21 (λDE3) enabling the expression of N-terminal His-tagged N-protein after induction with IPTG. The cells were growth in 100 ml LB medium at 120 rpm at 37°C to OD600nm of 0.4. The incubator temperature was lowered to 16°C, after 30 min, IPTG was added to a final concentration 0.3 mM and the culture was kept at 120 rpm at 16°C over/night. Cells were collected by centrifugation at 3,000 xg for 5 min. The cell pellet was resuspended in 20 ml of buffer 1 (Tris-HCl pH 8 50 mM, KCl 100 mM, glycerol 10%). Cells were disrupted by sonication on an ice bath. The soluble fraction was recovered after centrifugation at 20,000 xg for 20 minutes and loaded onto a 5 ml His-Trap column (Cytiva), which had been previously equilibrated with buffer 1. The column was washed with 10 ml of buffer 1 and bound proteins were eluted with buffer 1 containing increasing concentrations of imidazole. The His-tagged N-protein eluted at 300 mM imidazole. The final protein preparation yield was ∼ 8 mg and was homogeneous as judged by SDS-PAGE analysis. The purified protein was stored in aliquots at 4°C for one week or at -20°C for up to four months.

### Human samples

Samples were collected at the Hospital de Clínicas (CHC/UFPR), Hospital Erasto Gaertner, Hospital do Trabalhador and Secretária Municipal de Saúde de Guaratuba. Samples for serological analysis comprised both serum and plasma-EDTA. COVID-19 positive cases were confirmed by the detection of SARS-CoV-2 RNA by real-time RT-PCR from nasopharyngeal sample swabs. The time point of sampling of serum ranged from 1 to 70 days after onset of symptoms. The cohort of 210 negative controls consisted of pre pandemic samples. The Institutional Ethics Review Board of CHC/UFPR (n# 30578620.7.0000.0008), CEP/HEG (n# 31592620.4.1001.0098) and (CEP/HT n# 31650020.5.0000.5225) approved this study.

### ELISA assays

The purified SARS-CoV-2 N-protein was diluted to 2 ng/µl in 50 mM Tris-HCl pH 8, 100 mM KCl, 10% (v/v) glycerol and 0.1 ml aliquots were transferred to the wells of ELISA plates (Olen) and incubated overnight at 4°C. Unbound excess protein was removed, and wells were washed twice with 0.2 ml of 1x TBST buffer pH 7.6 (Tris 2.42 g.l^-1^, NaCl 8 g.l^-1^ and tween 80 0.1% v/v). Wells were blocked with 0.2 ml 3% (w/v) skimmed milk in 1x TBST overnight at 4°C. The serum was diluted at 1:1,000 in 1x TBST containing 1% (w/v) skimmed milk and 0.1 ml were loaded onto the ELISA duplicate wells following by incubation at room temperature for 1 hour. The wells were washed three times with 0.2 ml of 1x TBST followed by incubation with 0.1 ml of anti-human IgG-HPR from goat (Thermo Fisher - cat number 62-8420) diluted 1:3,000 in 1x TBST. Wells were washed three times with 0.2 ml of 1x TBST. The HPR substrate TMB (Thermo Fisher - cat number 00-2023) was added to the wells (0.1 ml). The reaction was stopped after 10 min incubation at room temperature by addition of 0.1 ml 1M HCl. The plates were put on the top of a white light transilluminator device and photographed. Samples were run in duplicates and data were reported as % of a positive reference serum. The optical density was measured at 450 nm using a TECAN M Nano plate reader (TECAN) monochromator at bandwidth 9 nm and 25 flashes.

The commercial Euroimmun Anti-SARS-CoV-2 ELISA IgG test was used following the manufacturer’s instructions.

### Data analysis

One COVID-19 positive serum was used as a positive control and reference throughout the study. All data were expressed as % of this positive control before applying Receiver Operating Analysis using GraphPad Prism 7.0. Statistical analyses were performed using the *t* test. To validate our ELISA assay to detect SARS-CoV-2 seroconversion in human a cohort consisting of 140 PCR-confirmed COVID-19 cases and 210 pre-pandemic negative controls were used. The pre-pandemic control sera produced only background signal which were clearly distinguished from the PCR-confirmed COVID-19 cases (Fig. 2A). The age of the negative cohort ranged from 21 to 86 years of age (mean = 62). It is likely that most of these subjects had experienced prior infections with common coronavirus (229E, NL63, OC43, and HKU which usually cause mild upper-respiratory tract disease, like the common cold). These data suggest that our ELISA assay shows negligible cross reactivity for IgG raised against common coronavirus. However, it is worth mentioning that we detected 4 reactive pre-pandemic samples whose signals were above the established cutoff thereby being classified as false positives (Fig. 2A). Such reactivity could be potentially explained by cross reaction with IgG raised against common coronavirus.

The ELISA method operated with an area under the ROC curve of 0.96 (Fig. 2B), the cut off the assay was set at 90% sensitivity and 98% specificity. These values are in the same range for other SARS-CoV-2 immunological IgG testes described previously (Cai *et al*., 2020; den Hartog *et al*., 2020; Rikhtegaran Tehrani *et al*., 2020). It is worth mentioning that among the PCR-confirmed COVID-19 cases, 32 samples were in the early stage of infection (≤ 10 days after PCR detection) and this may explain the low sensitivity. Indeed, if only samples collected ≤10 days after PCR detection were considered, the sensitivity dropped to 81%. It is also worth mentioning that 40 samples of the COVID-19 positive cohort were from asymptomatic/mild COVID-19 cases which are known to induce lower humoral response to SARS-CoV-2 infections (Long *et al*., 2020). Indeed, our analysis confirmed that the IgG titer was significantly lower in mild versus hospitalized COVID-19 patients (Fig. 2C).

A panel of 90 samples was analyzed in parallel by our developed ELISA and the Euroimmun Nucleocapsid-based IgG ELISA. All the 44 pre-pandemic negative samples analyzed were negative in both tests confirming the high specificity of the assays. Out of the 46 PCR confirmed COVID-19 samples, both testes missed 4 samples (false negatives). One sample was negative in both testes and three samples were uniquely negative in each of the two different tests. These data support that our ELISA performed with similar sensitivity to the commercial Euroimmun ELISA test.

In conclusion, here we describe a high yield and simple method to purify the SARS-CoV-2 Nucleocapsid antigen using only one simple affinity chromatography step. The antigen preparation was used to develop an indirect ELISA which allowed accurate detection of SARS-CoV-2 seroconversion in humans. The developed assay is ready for mass production in order to help to track COVID-19 cases specially in developing countries where access to molecular test is limited.

## Data Availability

I agree

## Acknowledgments

This work was supported by the Alexander von Humboldt foundation and The Federal University of Paraná, Brazil. MC received an undergraduate technological development fellowship from CNPq (PIBIT - UFPR).

## References

Cai, X., Chen, J., li Hu, J.-Long, Q., Deng, H., Liu, P., et al. (2020) A Peptide-Based Magnetic Chemiluminescence Enzyme Immunoassay for Serological Diagnosis of Coronavirus Disease 2019. J Infect Dis.

Cheng, V.C.C., Lau, S.K.P., Woo, P.C.Y., and Kwok, Y.Y. (2007) Severe acute respiratory syndrome coronavirus as an agent of emerging and reemerging infection. Clin Microbiol Rev.

F. Huergo,, L.A. Selim,, K.S. Conzentino,, M.C. M. Gerhardt,, E.R. S. Santos, A.Wagner, B., et al. (2021) Magnetic Bead-Based Immunoassay Allows Rapid, Inexpensive, and Quantitative Detection of Human SARS-CoV-2 Antibodies. ACS Sensors 0.

Hartog, G. den, Schepp, R.M., Kuijer, M., GeurtsvanKessel, C., Beek J. van, Rots, N., et al. (2020) SARS-CoV-2–Specific Antibody Detection for Seroepidemiology: A Multiplex Analysis Approach Accounting for Accurate Seroprevalence. J Infect Dis.

Huergo, L.F., Conzentino, M.S., Gerhardt, E.C.M., Santos, A.R.S., Pedrosa, F. de O., Souza, E.M., et al. (2020) Magnetic bead-based ELISA allow inexpensive, rapid and quantitative detection of human antibodies against SARS-CoV-2. medRxiv.

Long, Q.X., Tang, X.J., Shi, Q.L., Li, Q., Deng, H.J., Yuan, J., et al. (2020) Clinical and immunological assessment of asymptomatic SARS-CoV-2 infections. Nat Med.

Petherick, A. (2020) Developing antibody tests for SARS-CoV-2. Lancet.

Rikhtegaran Tehrani, Z., Saadat, S., Saleh, E., Ouyang, X., Constantine, N., DeVico, A.L., et al. (2020) Specificity and Performance of Nucleocapsid and Spike-based SARS-CoV-2 Serologic Assays. medRxiv.

Zhu, N., Zhang, D., Wang, W., Li, X., Yang, B., Song, J., et al. (2020) A novel coronavirus from patients with pneumonia in China, 2019. N Engl J Med.

